# Determinants of diphtheria outbreaks and predictors of outcomes; lessons learnt from 2025 outbreak in Imo State, Nigeria

**DOI:** 10.1101/2025.09.08.25335373

**Authors:** Adeniran Adeniyi Ayobami, Igboekwu Chukwumuanya, Wadzingi Williams Bassi, Eronini Ebubechi Chinwe, Ali Wada Aliyu, Okeji Austine Chidiebere, Evi-Parker Uloma, Egbuna Hyacinth Chukwuebuka, Gilbert Bestdone, Okoroama Chibuzo Linda, Ihemba Vivian Ngozi, Ihedioha Leonard, Izegbune Angela, Nosike Chibuzo Job, Nnawuba Victor, Ekeoba Chiamaka Cynthia, Uche Chukwudi

## Abstract

Diphtheria is a potentially fatal infectious that presents mostly with respiratory symptoms in which an exotoxin produced by the causative pathogen causes tissue destruction and death if untreated. This quantitative, descriptive, longitudinal study aims to determine the determinants of diphtheria outbreak and the predictors of outcomes in Imo State, in southeast Nigeria.

Data was collected during the outbreak response using standardized Nigeria Centre for Disease Control, NCDC, data tools deployed for the response, entered into an Excel ® 2016 line-list, visualized and exported into SPSS® version 20 software for analysis. Additional immunization and surveillance data were extracted from the DHIS2 and SORMAS respectively. Analysis was done using descriptive statistics and associations between categorical variables were tested using Fisher’s exact method at 0.05 level of significance. A total of 205 suspected and 160 confirmed cases of diphtheria were reported across 73 wards in 17 LGAs of Imo State. Among these, 160 were confirmed cases; 67 laboratory-confirmed cases, 11 epidemiological linkages, and 82 were clinically compatible. A total of 15 deaths were recorded with a case fatality rate (CFR) of 9.4%.

There were more females (110, 53.7%) than males (95, 46.3%), 73% (n=11) of deaths were among the males while 27% (n=4) were females. The study found a significant association between gender and death from the infection (p < 0.05). The infection affected predominantly children with 74.6% (n = 153) affecting ages 0 – 14 years and 93.4% (n = 192) occurring below the age of 40. All deaths (100%, n = 15), occurred among children between 4-9 years, late presenters (78%), children with bullneck (100%) and children with no DAT administered at the point of admission (93.3%).

## Introduction/Background

Diphtheria is a potentially fatal infectious disease caused by toxin-producing strain of Corynebacterium diphtheriae (C. diphtheriae) and rarely by toxin-producing strains of C. ulcerans and C. pseudotuberculosis. The C. diphtheriae typically has an incubation period of 2 to 5 days, though it can extend up to 10 days.

The disease presents mostly with respiratory symptoms; in which case, the exotoxin produced by the toxin-producing strains of the pathogen causes necrotic destruction of tissues of the upper airway leading to the formation of greyish-white patches referred to as pseudomembranes in the upper respiratory tract [1]. This toxin also spreads to other organs like the myocardium and peripheral nerves causing acute obstructive symptoms, acute systemic toxicity, myocarditis and neurologic complications [2]. These systemic manifestations are due to inhibition of protein synthesis, cell death and necrotic tissue destruction caused by the exotoxin and are associated with increased risk of death [3].

Although uncommon, diphtheria infection also affects the skin, mucous membranes and the genitalia [2]. Transmission is by intimate respiratory and direct contact from person to person except for cutaneous diphtheria infection, which is usually zoonotic. The incubation period of diphtheria infection lasts between one to ten days, if untreated however, infected person may remain infectious for as long as six weeks [2]. Prompt treatment with effective antibiotics, within a period of 48 hours, has been found to reduce infectivity. Case fatalities during outbreaks could be as high as 10 percent, especially in areas where diphtheria antitoxin, which is mainstay of treatment, is not readily available [2].

Infected individuals develop fever, sore throat, difficulty with swallowing, drooling of saliva in children and neck swelling, with enlargement of lymph nodes in the neck; typically described as ‘bull-neck’. This is followed by the development of the pseudomembranous, thick, greyish patches over-laying the tonsils [5].

If left untreated, the diphtheria exotoxin begins to invade major organs, leading to damage to the nervous system, renal complications and inflammation of the heart myocardium where it becomes fatal even with treatment [8, 9].

Laboratory diagnosis of the diseases is usually done to validate clinical diagnosis as most cases can be correctly diagnosed clinically. It is typically done through culture aimed at isolating the Corynebacterium diphtheriae, followed by tests for the detection of toxin production [8, 9]. Vaccination programs have significantly reduced the global incidence of diphtheria. Outbreaks, however, still occur, especially, in areas of poor vaccination coverage or when immunization wanes as population ages. In the 1990s, the Russian Federation and the newly independent States of the former Soviet Union reported more than 157,000 cases and 5,000 deaths of diphtheria [8, 9]. Also, since 2016, there has been a steady rise in the number of cases reported worldwide, which doubled from an average of 8105 cases between 1997 and 2017 to 16651 in 2018 [6].

In Nigeria, outbreaks of diphtheria are beginning to re-emerge and about 43,743 suspected cases have been reported across the 37 States in 360 of 777 local government areas as of 3rd May 2025 [9]. Seven States, Kano; 24,415, Yobe; 5,330; Katsina; 4,355; Bauchi; 3,066; Borno; 3,064, Kaduna; 840 and Jigawa; 364, accounted for 96% of the suspected cases reported. With a test positivity rate of 60.6 percent, a total of 1376 deaths have been reported from 26,499 confirmed cases giving a case fatality rate of 5.2 percent [10]. A major outbreak which occurred in Kano State, Northwest Nigeria, has been declared as the worst in a decade. The outbreak, which started which started in May 2022 reported over 18,320 cases with over 800 deaths and a case fatality rate (CFR) of 4.5%. The study reinforces previous reports of the weak health systems worsened by the covid-19 pandemic [11].

**Figure 2:**
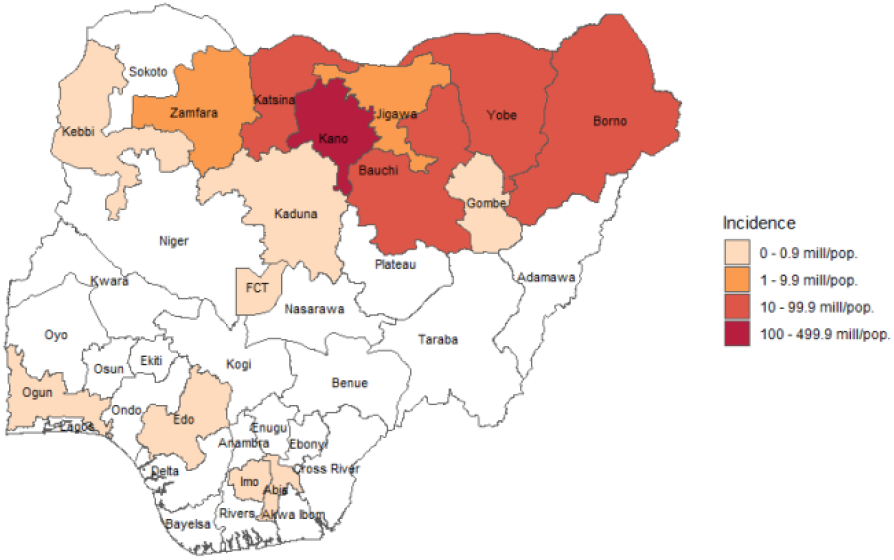
Incidence (per million population) of confirmed diphtheria cases in Nigeria by States, epi-week 19, 2022 to epi-week 18, 2025. Source: NCDC

Diphtheria is both preventable and treatable. Treatment outcomes are however dependent on several factors like the age, timeliness of presentation and the availability of diphtheria anti toxin. Pentavalent vaccine is a combination vaccine with five individual antigens conjugated into one. This five-in-one vaccine protects against diphtheria, tetanus, whooping cough, hepatitis B, and Hemophilus influenzae type B, and it is generally used in middle- and low-income countries, where polio vaccine is given separately [4]. In Nigeria, children under the age of five can get vaccinated against the disease with these vaccines. They can also be given the diphtheria-tetanus toxoid combination (TD) vaccines, which only fight against tetanus and diphtheria [10, 12, 13]. Another combination is the tetanus, diphtheria, and pertussis (DTP) vaccine for adults and older adolescents, which can also be administered to pregnant women [10, 12], although the DPT has now been replaced by Pentavalent vaccine in Nigeria’s routine immunization schedule.

The treatment of diphtheria usually takes a multi-disciplinary approach involving various specialties such as, pediatrician, infectious disease specialist, ENT specialist, cardiologist, pulmonologist, nephrologist, dietician, intensive care physician and nurses. Management involves triage and resuscitation, isolation, definitive therapy with the use of appropriate antibiotics and diphtheria antitoxin, supportive therapy, management of complications and management of contacts [14].

Complications following delayed treatment include the bull neck appearance which comes as a result of cervical adenopathy and swollen mucosa. This, along with aspiration of the pseudo-membrane also contributes to airway obstruction or suffocation. Within 1-12 weeks, infected patients develop myocarditis, congestive heart failure, conduction abnormalities, arrhythmias, debilitating neurologic dysfunction and renal failure [14].

Several studies have been conducted to determine the specific determinants of diphtheria outbreaks, and the specific predictors of outcomes, in populations. Asides that before the development of vaccines, the disease was more common in young children, studies conducted in Pakistan found that and that lack of requisite trainings and experience among health workers account for most late referrals at irreversible stages of the disease [14]. Weak immunization programs and poor hygiene have also contributed to outbreaks in this south Asian country [5,14, 15]. The WHO recommended that to ensure long-term protection, diphtheria toxoid should be administered to infants as a primary series of three doses, followed by three appropriately spaced booster doses [2,3].

In Nigeria, outbreaks have occurred across the country, including in areas with high vaccination coverage. Studies have demonstrated the risk of re-emergence, and outbreaks, of diphtheria in countries where the recommended vaccination programmes are not sustained, and increasing proportions of adults are becoming susceptible to diphtheria [3].

This study aims to determine the determinants of diphtheria outbreaks in Nigeria from the lessons learnt following the 2025 outbreak in Imo State, Nigeria.

## Methods

This quantitative, descriptive, longitudinal study was conducted in Imo State, Southeastern Nigeria. The State, with a projected population of about 7.2 million (2025 projection of National Population Census, 2006) has twenty-seven Local Government Areas (LGA) with four hundred and eighteen wards. The population of children under five years of age is estimated at about 1,431,785 (20% of the total population), estimated population under one year of age is at about 286,357 and estimated population of women of reproductive age is 1,478,806. Residents are of homogenous Igbo ethnicity.

Data was collected during the outbreak response through the LGA disease surveillance and notification officers and from the designated diphtheria treatment centers. Data tools deployed were the standardized, NCDC approved case investigation forms (CIFs). Data from these CIFs were entered into an Excel ® 2016 line-list. Data was visualized and exported into SPSS® version 20 software for analysis. Additional immunization and surveillance data were extracted from the DHIS2 and SORMAS respectively. Analysis was done using descriptive statistics and associations between categorical variables were tested using Fisher’s exact method at 0.05 level of significance.

## Results

A total of 205 suspected and 160 confirmed cases of diphtheria were reported and line-listed during the outbreak that lasted between 3rd May 2025 and 31st July 2025. The cases were reported across 73 wards in 17 LGAs. Among these, 160 were confirmed cases; 67 laboratory-confirmed cases, 11 epidemiological linkages, and 82 were clinically compatible. A total of 15 deaths were recorded with a case fatality rate (CFR) of 9.4%.

Seventeen (17) of 27 LGAs (63%) reported at least, one suspected case with Aboh Mbaise LGA, where the index case was reported, reporting the highest number of cases, 114 (55.6%) followed by Owerri North LGA which is contiguous with Aboh Mbaise LGA, 26 cases (12.7%). Eight contiguous LGAs, Aboh Mbaise, Owerri North, Orlu, Owerri Municipal, Owerri West, Ezinihitte Mbaise, Ngor Okpala, and Ahiazu Mbaise accounted for over 90% of the reported cases (See Table 1 below). Also, Aboh Mbaise LGA reported 80% (n = 12) of the mortalities with a CFR of 10.5%.

**Table 1:**
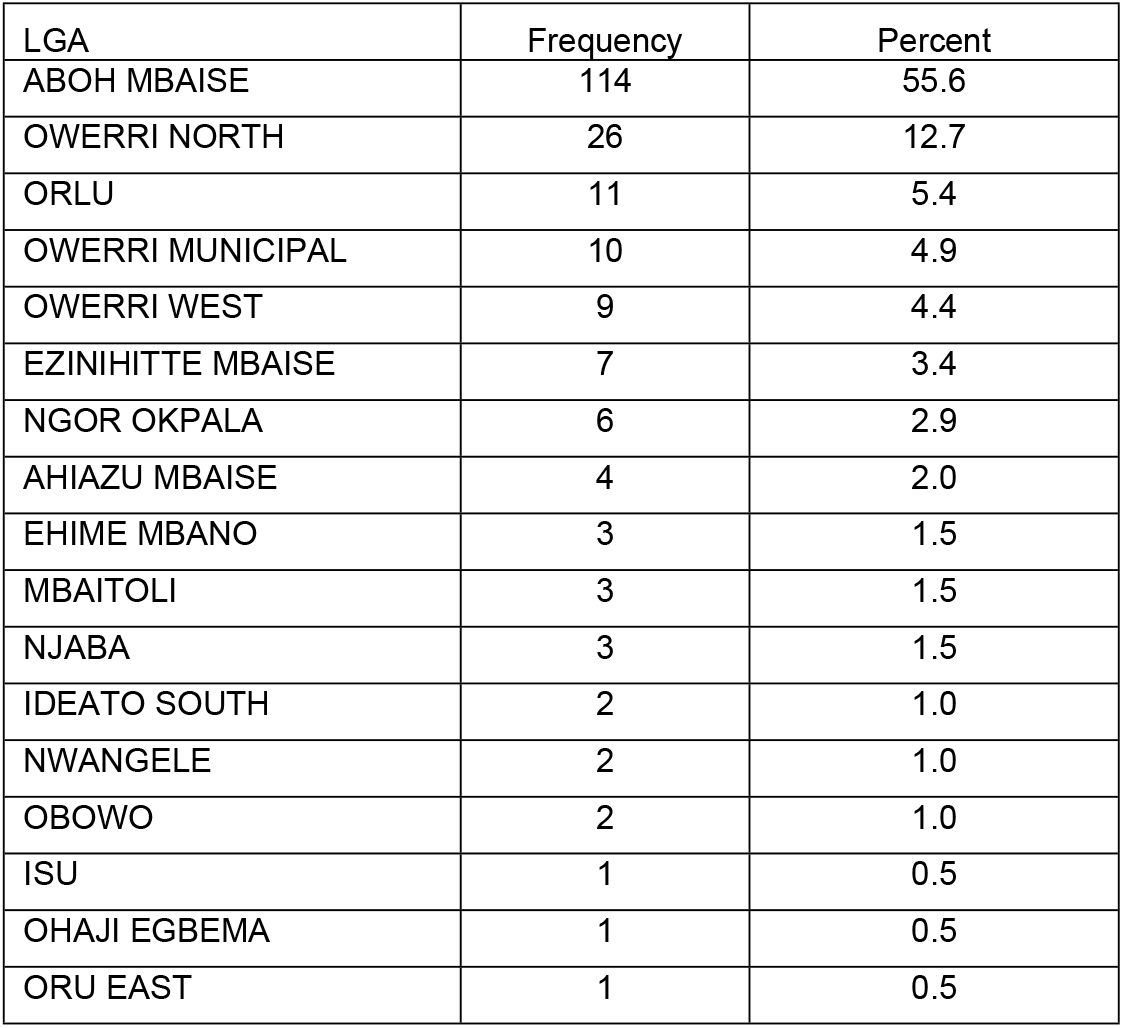
Distribution of reported cases of suspected diphtheria across LGAs.

| LGA | Frequency | Percent |
| --- | --- | --- |
| ABOH MBAISE | 114 | 55.6 |
| OWERRI NORTH | 26 | 12.7 |
| ORLU | 11 | 5.4 |
| OWERRI MUNICIPAL | 10 | 4.9 |
| OWERRI WEST | 9 | 4.4 |
| EZINIHITTE MBAISE | 7 | 3.4 |
| NGOR OKPALA | 6 | 2.9 |
| AHIAZU MBAISE | 4 | 2.0 |
| EHIME MBANO | 3 | 1.5 |
| MBAITOLI | 3 | 1.5 |
| NJABA | 3 | 1.5 |
| IDEATO SOUTH | 2 | 1.0 |
| NWANGELE | 2 | 1.0 |
| OBOWO | 2 | 1.0 |
| ISU | 1 | 0.5 |
| OHAJI EGBEMA | 1 | 0.5 |
| ORU EAST | 1 | 0.5 |

Out of the 205 suspected cases, there were more females than males (Male 95, 46.3%; females 110, 53.7%. Also 55%, (n=36) of laboratory confirmed cases were females while 45% (n=30) were males. However, 73% (n=11) of deaths were among the males while 27% (n=4) were females. Although there was no significant association between gender and diphtheria infection (p > 0.05), the study found a significant association between gender and death from the infection (p < 0.05).

The infection affected predominantly children with 74.6% (n = 153) affecting ages 0 – 14 years and 93.4% (n = 192) occurring below the age of 40. All deaths (100%, n = 15), however, occurred among children between 4-9 years. Among 64 patients who presented early at the health (within 48 hours), 78% (n = 50) were children between 0 −14; and 87% (n = 13) of the deaths occurred among children who presented late at the health facilities (greater than 48 hours) and before sample collection.

Fifty-two of the confirmed cases were managed as in-patients at the diphtheria treatment centres out of which 50 (96.2%) were discharged and 2 (3.8%) deaths recorded. All 15 deaths (100%) recorded presented with bullneck, 14 (93.3%) did not receive DAT at the point of admission, 4 (26.7%) presented with pseudo-membrane, 4 (26.7%) were hospitalized.

Out of the 33 patients recorded to have received DAT at the point of presentation at the health facility, 97% (n = 32) recovered with 3% (n = 1) death. Early administration of DAT was significantly associated with the chances of survival (p < 0.05).

Data extracted from the health facilities immunization data shows that as at the end of 2024, Mbutu 1 ward of Aboh Mbaise, the epi-center of the outbreak had a pentavalent coverage of 49% and about 135 unimmunized children (See Table 2 below). The affected wards are, also, all fully accessible and there are no special vulnerable populations.

**Table 2:**
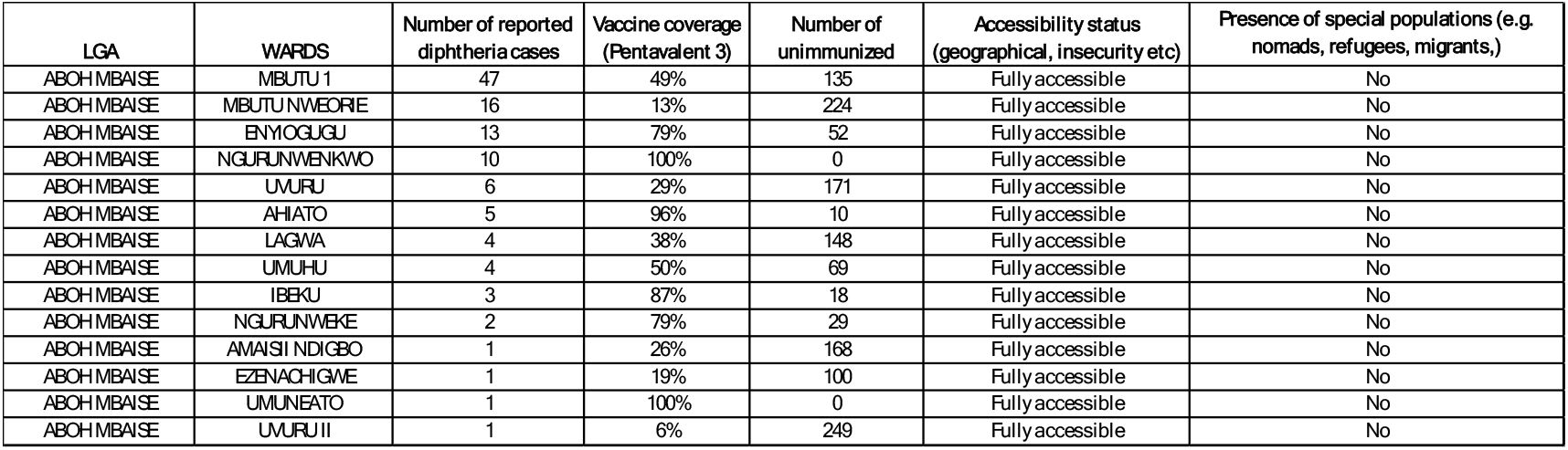
Vaccine coverage and other risk assessment status of the affected wards at Aboh Mbaise LGA as at the end of 2024. Source: WHO Polio risk assessment.

## Discussion

Several studies have described the determinants and drivers of diphtheria outbreaks in Nigeria. The World Health Organization, and the Office for the Coordination of Humanitarian Affairs (OCHA) in Nigeria reported that children and people who have received only a single dose of diphtheria vaccine, crowded housing and unhygienic areas, occupational exposure to suspected or confirmed diphtheria cases, and poor access to diphtheria antitoxin are major drivers of diphtheria outbreaks in Nigeria [15, 16, 17, 18, 19]. This is in keeping with the findings of this study which shows over three-quarters of affected people, and all deaths, occurred among children who met one or more of the above characteristics.

A study conducted in 2025 by Linda, O.C., et al, on pentavalent vaccine coverage in Imo State reported a rapid decline in vaccine coverage from 2020 to 2021 during the COVID-19 pandemic and a continuous decline in the State vaccine coverage till 2023 [4]. Aboh Mbaise LGA had persistently low pentavalent coverage over the six-year period [4]. Linda, et al, predicted the possibility of re-emergence of vaccine preventable diseases in the areas of low vaccine coverages [4]. During this 2025 outbreak, which is the focus of this study, Aboh Mbaise reported both the index case and the highest number of cases and deaths of the outbreak, at (114, 55.6%) and (12, 80%) respectively. This portrays the importance of vaccination and herd immunity as a key determinant of outbreak among this populations.

While there were more suspected and confirmed cases among the females (53.7% and 55% respectively), more deaths were reported among the males (73%). All these deaths occurred among children, hence, could not have been attributed to documented poor health seeking practices among the male folks [25, 26]. Ibrahim O.R., et al., (2022) in a similar study conducted following a diphtheria outbreak in Katsina, Northwestern Nigeria documented that age, sex, and even immunization status were not predictive of deaths from diphtheria infection among the 24 deaths recorded [26]. Thus, this observation in this study, might be attributable to the very small sample size of 15 deaths. This study also observed that majority of the mortalities (87%) were among children who presented late at the health facilities (greater than 48 hours) and before sample collection.

Mortality rates among the cases that were managed as in-patients at the diphtheria treatment centers (DTCs) was lower (3,8%) than the total CFR of 9.4%. Major predictors of mortality were presence of bullneck (100%) and non-administration of DAT within 24 hours of presentation (93.3%). This aligns with the finding of Ibrahim O.R., et al (2022) in Katsina State, which attributed high mortality among children to delayed presentation and unavailability of diphtheria antitoxin (DAT) which is the main stay of diphtheria treatment [2].

Although the outbreak was still ongoing as that the time of data collection, the case fatality rate recorded so far (9.4%) is however significantly lower than that recorded in Katsina (68.6%), Benin, Edo State (33.3%) [28] higher that reported in Kano (4.5%) [28, 29] and also higher than some low-middle income countries such as Indonesia (3.5%) [29], Bangladesh (0.9%),[31] and India (2.2%) [30].

## Conclusion

This study demonstrated the importance of addressing the deep-rooted causes of poor vaccine uptake, poor coverages, and low herd immunity in Imo State Nigeria. Stock-piling, pre-positioning and availability of DAT at designated treatment centers at the sub-national level should also be considered by the government and disease control agencies in Nigeria. The study suggests that the presence of bull neck in every child must be considered a major criterion for intensive and multi-specialized care at the DTCs in order to reduce mortalities from the infection.

## Data Availability

All data used for this study are available on the SORMAS and DHIS2 data platforms

